# COVID-19 in meat plants: activation of a Target Prevention Plan, in Italy

**DOI:** 10.1101/2023.05.08.23289661

**Authors:** Giorgio Di Leone, Luigi Bertinato, Gianfranco Brambilla, Valerio Manno, Flavio Napolano, Simona Savi, Gaetano Settimo, Domenico Lagravinese

## Abstract

During the COVID-19 pandemics, several outbreaks have been recorded all other the world in industrial slaughterhouses and meat processing plants. Occupational preventive medicine in such non-healthcare frontline essential services accounts for combined different environmental, social, and economic factors, to reduce the burden of COVID-19 in the workplaces and in the connected residential settings. In Italy, during the first year of the pandemics, an advocacy action has been activated, targeted on meat plant managers and related food business operators. A risk-oriented control plan was agreed by competent Italian Health Authorities at Region/Province level. A questionnaire focused on the inventoried risk factors reported in the literature in such working places have been developped as supporting tool, and administered on voluntary basis to the interested stakeholders. In addition, an outbreak questionnaire was proposed to the Prevention Depts of the Local Health Units. In the 2021 – 2022 years timeframe, we collected 333 advocacy and 24 outbreak questionnaires, respectively, on 4,765 inventoried plants at national level. Responses came mainly from those districts that locally activated the risk-oriented control plan. The lack of awareness to update the Risk Assessment Document of the meat plant for COVID-19, non instrumental body Temperature checks of workers at the entrance, working force from different subcontractors, poor hygiene in the shared places and insufficient ventilation represented the main critical points recorded. The cross-checks between the results from the advocacy and from the outbreak questionnaires are feeding an after-action review for such food-chain related essential work settings within a One Health approach.

## 1. Introduction

During the Covid-19 pandemics, it became progressively clear that also some occupational non healthcare settings could be vulnerable to outbreaks: this was the case of meat plants (slaughterhouses, meat processing and cutting plants) where the combination of environmental, social, and working condition factors represented a driver for contagiousness among workers [1], extended in some cases to relatives in the residential settings [2]. (Taylor et al., 2020). From 2020, rapid and large-scale COVID-19 outbreaks in high throughput industrial meat plants with a working force up to 20,000 workers have been reported in the US and Canada, and in the European Union [3-5]. The epidemiological investigations have more and more elucidated the combined risk factors that favour the Sars-CoV-2 enter and persistence in the working places, and the spread among workers, such as: a) Workforce recruitment and turnover, collective transport systems to/from the working place, and housing [6-8]; b) Working places with poor ventilation and insufficient fresh air exchange; c) Presence of aereosol/vapours able to transport the virus well above the 1-2 m distance prescribed among workers, and d) cool surfaces where virus particles could condense and persist for days [9]. COVID-19 and more in general contagious infectious diseases outbreaks in such essential settings for the food chain represent a food insecurity factor, as matter industrial slaughterouses are a critical point to guarantee the meat supply chain from farm to fork: if the abatement of intensive farmed high inbred poultry and pigs is postponed only for few days, animal welfare and meat quality worsten, thus hampering the cost effectiveness of the food production and provoking food waste. In addition, the presence of Sars-Cov-2 genome on the surface of packaged meat [10, 11] has been a matter of international import/export dispute within the Word Trade Organization [12], thus causing food chain disruption.

In this paper we aim to describe the results of a target plan on COVID-19 prevention in meat plants in Italy, set up at the end of 2020. Such target prevention plan, designed on the basis of the first epidemiological evidences in Italy and abroad, has been proposed to the Regional and Province Health Authorities for its adoption and implementation, despite the urgent priorities in the healthcare territory setting determined by the evolution of the pandemics in Italy. **Figure 1** illustrates the activation steps of this plan in the context of the recorded pandemic curve in healthcare and non-healthcare workers in Italy from official data (https://www.epicentro.iss.it/coronavirus/sars-cov-2-dashboard).

**Figure 1.**
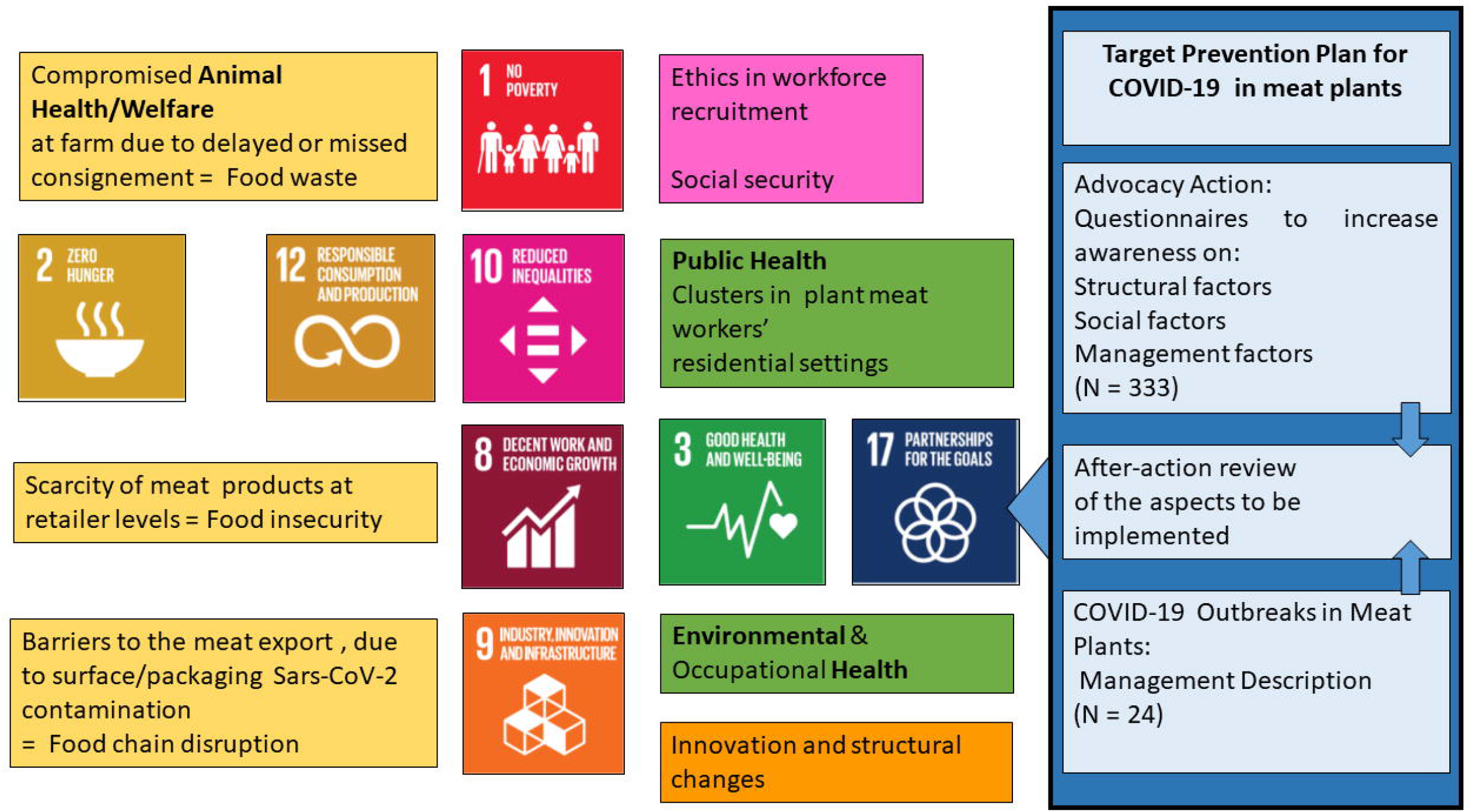
COVID-19 pandemic curve in Italian workers, along with the events leading to the activation of a Target Prevention Plan in Meat Plants.

## 2. Materials and Methods

### 2.1 Set up of questionnaires for COVID-19 Occupational Health in meat plants

In Italy, the activity of preventive medicine in occupational settings has been assigned to Regions and Provinces, under the co-ordination of Italian Regions Conference, technically supported by the Working Group on “Health and Safety at the working places”. This WG has mandate to propose the activation of the so called “Target Prevention Plans” to the Regions and Provinces; these plans consist on an advocacy action with the stakeholders to reach priority and risk-oriented targets of health prevention in the working places. Within this frame, on April 2020 the Istituto Superiore di Sanità, as Technical Scientific Institution of the Italian Health System contacted the Local Health Unit of Bari – Apulian Region, who reported the first COVID-19 outbreak in a meat plant [13]; on the basis of the evidences from the field and those reported in the scientific literature, an advocacy webinar was organised on September 2020, with the involvement of meat plants associations, national, regional, province authorities, to share evidences and experiences. The final outcome was the proposition of three different tools to support a target COVID-19 prevention plan in meat plants: 1) a first questionnaire addressed to meat plants management about the critical control points to be considered for the prevention and risk assessment of COVID-19; 2) a second questionnaire addressed to local health authorities for the reporting of COVID-19 outbreaks; 3) a harmonised check list for the official inspection by competent local authorities at meat plants. The first two questionnaires, after a first on-field validation, have been published with an Italian and English versions in the COVID-19 Reports edited by Istituto Superiore di Sanità [14], and made freely available to stakeholders via Google Modules platform, under the General Rules for Data Protection. Italian Meat Plant Associations were asked to inform their members about the opportunity participate to such survey on voluntary basis. The responses to the advocacy and the outbreak questionnaires were collected from the end of December 2020, till the end of December 2022, and reported as results in this paper. In **Figure 2** the flow-diagram illustrates the main health-related stakeholders and the COVID-19 prevention activities at the working places

**Figure 2.**
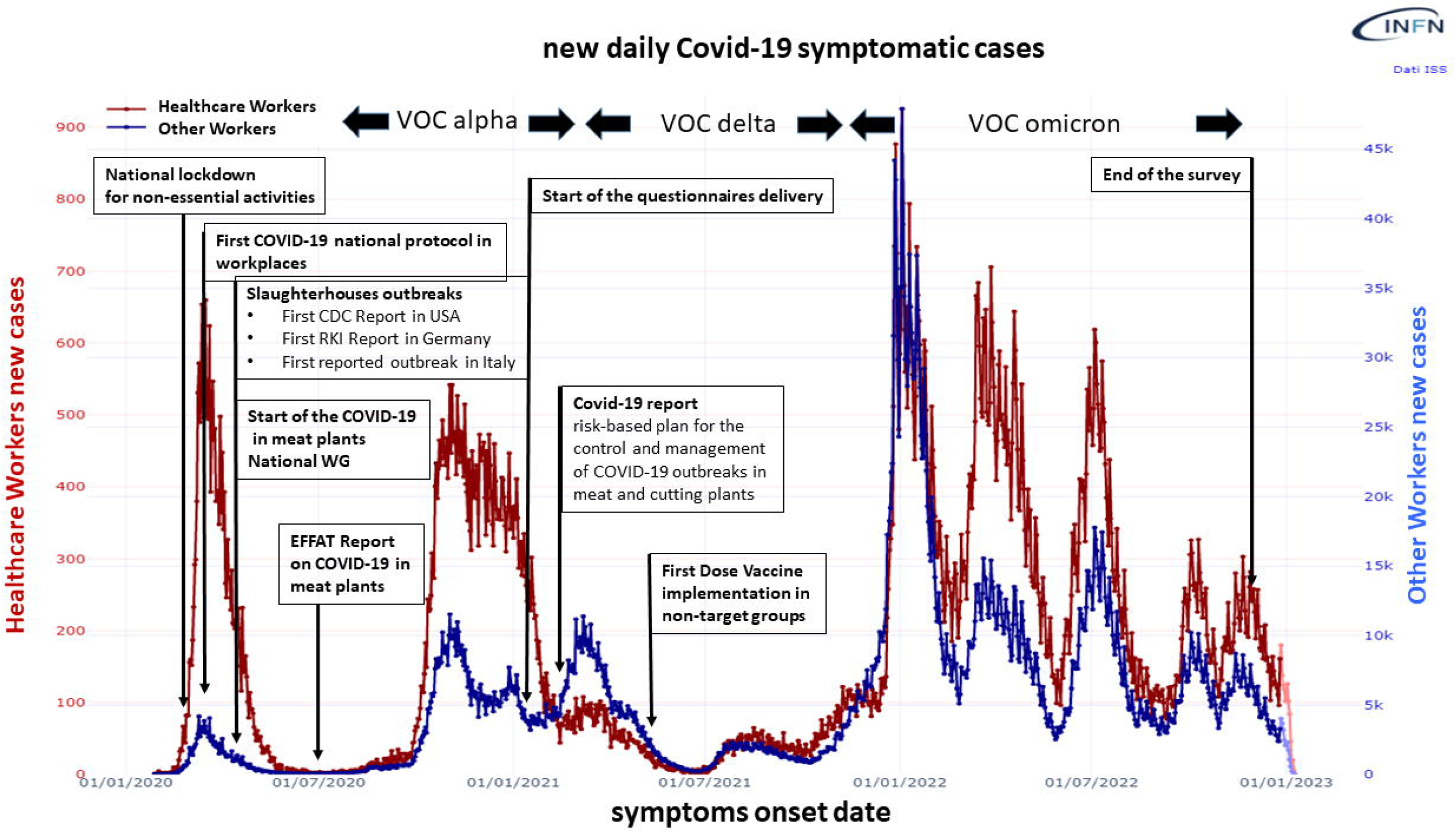
Flow diagram of the stakeholders (boxed) and the activities (circled) involved in the set up of a COVID-19 risk-oriented prevention plan in Meat Plants, in Italy.

## 3. Results

### 3.1 Awareness about the critical control points to be considered by meat plant management

#### 3.1.1 Meat plant profiling and workforce

During the considered two years timeframe (20-22), we recorded 333 filled and validated COVID-19 awareness questionnaires on a total of 4,675 slaughterhouses and meat cutting and processing plants inventoried at central level by the competent Authority (https://www.dati.salute.gov.it/dati/dettaglioDataset.jsp?menu=dati&idPag=8).

The geographical provenience of such modules acknowledges in large part those Italian Regions/Autonomous Provinces who explicitly declared their interest in such Targeted Prevention Plan. In the details: Lombardy (N =140), Veneto (N=95), Trentino (N =37), Calabria (N=29), Apulian (N= 11), Piedmont (N=9), Emilia-Romagna (N=6), Sardinia (N=3), Lazio, Sicily and Umbria (N= 1), respectively.

In **Table 1**, we report the different plant tipologies, that in some cases acknowledge the presence of a cutting plant associated to the slaughterhouse, and activities addressed to different animal species.

**Table 1:** Profile of the slaughterhouses and meat processing/cutting plants that participated to the questionnaire on voluntary basis. Due to the presence in the same plants of activities addressed to different animal species, the total number reported in Table 1 is greater than the number of the recorded questionnaires (N = 333).

| Species | Activities |  |  | Total |
| --- | --- | --- | --- | --- |
|  | S | PC | S+PC |  |
| cattle | 104 | 46 | 18 | 168 |
| horses | 44 | 6 | 5 | 55 |
| cattle and horses | 106 | 48 | 18 | 172 |
| pigs | 131 | 85 | 18 | 234 |
| small ruminants | 72 | 12 | 10 | 94 |
| poultry | 27 | 18 | 4 | 49 |
| rabbits | 9 | 9 | 1 | 19 |
| poultry and rabbits | 30 | 22 | 5 | 57 |

S = slaughterhouse, PC = processing/cutting plant; S+PC = slaughterhouse and processing/cutting plant.

Most of the responding plants indicate a working activity not extended on all the 6-7 week days. This implies the working force mostly is not shifted within the same working day (N = 302; 91%). Two and three working shifts/day have been recorded in the 5% and 2% of the plants respectively. Missed answers = 2%. Full details in **Table 2a**.

**Table 2.** a) Working days per week (left) and b) Number of Workers in those plants who participated to the survey (right).

| Working days | S | CP | S+CP | Total | Workers | Plants |
| --- | --- | --- | --- | --- | --- | --- |
| 1 | 54 | 8 | 4 | 66 | N | N |
| 2 | 61 | 7 | 4 | 72 | 1-5 | 105 |
| 3 | 15 | 8 | 2 | 25 | 6-10 | 52 |
| 4 | 14 | 7 | 3 | 24 | 11-25 | 55 |
| 5 | 45 | 60 | 7 | 112 | 26-50 | 31 |
| 6 | 13 | 13 | 7 | 33 | 51-100 | 15 |
| 7 | 1 | - | - | 1 | 100+ | 42 |
| Total | 203 | 103 | 27 | 333 | missed | 33 |

3 S = slaughterhouse, PC = processing/cutting plant; S+PC = 4 slaughterhouse and processing/cutting plant

Non permanent staff (cooperatives, third parties, autonomous workers) is present in the 42% of the companies (N = 139). Cooperatives are regularly present in 125 plants: 52/125 plant managers engage one cooperative; 36/125, two; 18/125, three; 10/125 four-five; 9/125, 6 or more. The working force from non permanent staff is generally shared over different activities, from livestock handling, to cleaning and packaging. The overall number of workers (permanent, non permanent) present on average in the responding plants is reported in the **Table 2b**.

#### 3.1.2 Preventive measures at the working place and personnel management

All the responding food business operators declare workers have been properly informed about the preventive measures to be taken in case of suspect of Covid-19 (such as: stay at home, if symptomatic; call the appointed physician of the healthcare system; alert the plant staff if symptomatic at the working place and avoid close contacts; follow the rules to prevent contagiousness at the working places. The regular instrumental check of the body Temperature at the entrance of the plant has been reported in the 73% of answers (243/333); in 56 cases (17%) it was asked a self-declaration, and in 1 case, this procedure is omitted. Missed/inconsistent answers were notably the 10%. Independent and time-shifted way in/way out pathways for workers and visitors are present in 305/333 cases (92%), while dedicated toilets only in the 37% of cases (123/333).

Visitors are not informed about preventive measures in 19/333 cases (9%), and appropriate checks about the appropriate and regular application of the preventive measures not fully implemented in 123/333 plants (37%). Regular cleaning and sanitization of the shared places, changing rooms, canteens are in place in the 95% of the plants, with daily frequencies in the 59% of cases, three/four times a week in the 15%, and once/twice a week in the 15%, respectively. The remaining 1% report a cleaning and sanitization interval over 7 days.

The accessible and easy-to-find presence of hand-washing dispensers is declared in the 99% of the prevention questionnaires. Personal Protective Equipments (PPEs) declared always available in 278/333 (83%) of cases, when interpersonnel distance are less than 1 meter, according to the national guide-lines issued by the Italian Government on March 20 (Decreto del Presidente del Consiglio dei Ministri 11 marzo 2020), and the technical uptated on April 20 by the National Institute for Insurance against Accidents at Work, a public non-profit entity safeguarding workers against physical injuries and occupational diseases (https://www.inail.it/cs/internet/docs/alg-pubbl-rimodulazione-contenimento-covid19-sicurezza-lavoro.pdf). In 16 cases (5), because a workers’ distance above 1 m, the answer is negative. Missed/wrong answers accounted for 10%. PPE were reported to be changed every day in the 99% of cases, and information and instruction about their their proper use/wearing present in 315/333 answers (95%). Details about the PPE used in the plants, according to the working force were present in 293 questionnaires, and are reported in **Table 3**.

**Table 3.**
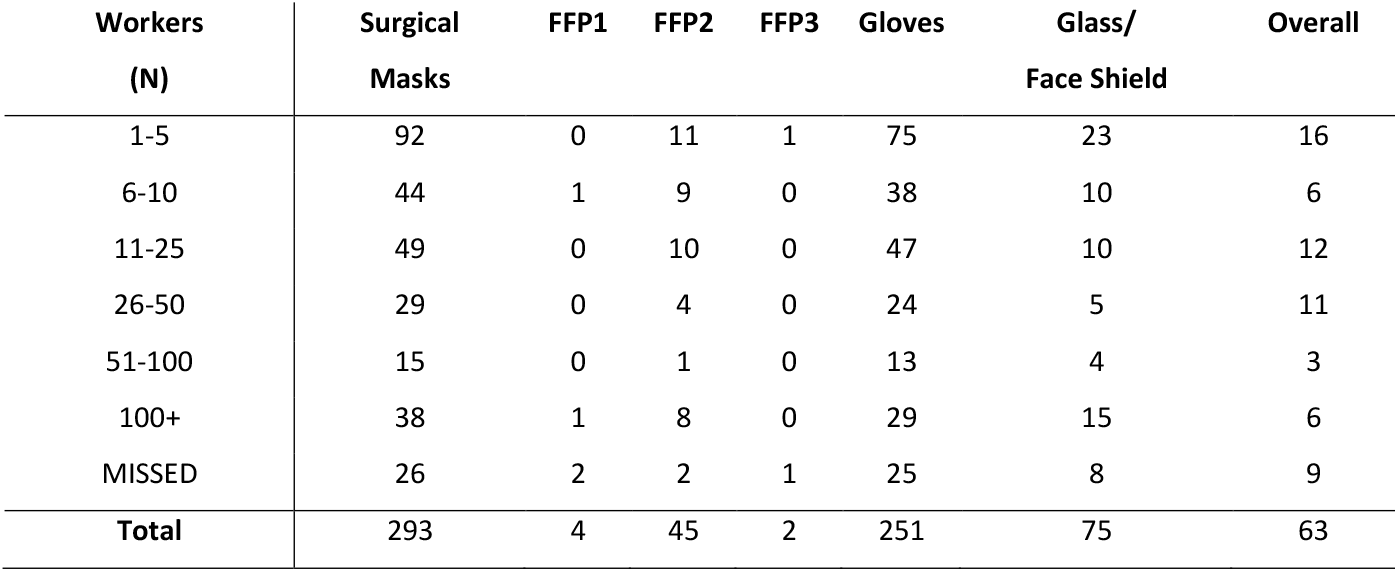
Profile of PPE availability in each of the 293 responding plants, according to the number of workers

Appropriate workplace organization to keep the prescribed minimum distance of 1 meter between workers has been implemented in 173/333 plants (53%); in 135 cases (41%) it has been reported the implementation of such measure is not necessary. The remaining 6% of plants gives negative or missed answers. The presence of physical barriers between workers has been reported in 116 answers (35%), while in the 50% of cases, it is declared the absence of a specific need.

The time zones at the entrance and exit have been designed to respect the 1 m distance in 168 plants (50%), while in 143 it is not the case. Negative answers account for the 7% (N=23). Staggered shifts to the canteen and to shared places are present in the 87% of cases (N=290).

Smart working is implemented in 63 plants)19%) for those non-essential activities; a personnel turn-over plan to reduce the contacts has been reported in 71 answers (21%), with negative answers in the remaining 263 cases. Social valves in case of absence from the working place due to Covid-19 illness have been activated in 86 plants (26%). In 164 cases (49%) workers have been asked to take holidays, with negative and missed answers in the remaining 25%.

The activation of a committee (with occupational health responsibles and trade union representatives) (**Figure 2**) in charge to verify the application of the COVID-19 preventive measures has been reported in 171 plants (51%), and the COVID-19 update of the mandatory document on the risk assessment in the 83% of answers (83%).

#### 3.1.3 Ventilation and vapour/areosol formation

In **Table 4** we report the recorder anwers for the questions related to the ventilation, according to the different plant premises. The regular maintainance of HVAC filters is declared in the 90% of plants, while records of ventilation mantainance is reported in the 50% of the plants, only.

**Table 4.** Number of natural ventilation vs mechanical Heating, Ventilation, Air Conditioning (HVAC) in the premises of 333 meat plants: between (brackets), the number of premises with HVAC air recycling > 30%.

| Premise | Ventilation |  |  | Total |
| --- | --- | --- | --- | --- |
|  | Natural | HVAC | HVAC<br>Not Reported |  |
| Working area | 234 | 91 (26) | 8 | 333 |
| Offices | 306 | 8 (2) | 19 | 333 |
| Changing Room | 288 | 25 (7) | 20 | 333 |
| Canteen | 156 | 17 (6) | 160 | 333 |
| Shared Places | 245 | 24 (7) | 64 | 333 |

#### 3.1.4. Vapours and aerosols generation

In 287 plants (86%) the cleaning of the working places acknowledges the use of high pressure water jets. Among them, 65 plants (20%) report such operation is carried out in presence of workers. In pig slaughterhouses, scraping is performed with hot water baths (N=94), with steam and water (N=7), with singeing (N= 52), with brushing and showering (N=43), with infrared beams (N7). Missed answers account for N=33. In those scraping activities where water is used, 80 answers report such procedure in presence of workers. The distance from the vapour/aerosol source is <1 m in 7 plants, >1>2m in 38, and >2 in 35, respectively. Aspiration systems are in place in 59/89 plants. In poultry slaughterhouses the use of water for animal electric stunning as proxy for vapour generation is reported in 23/32 plants. Nebulizer systems to improve animal welfare in the pre-slaughter area and lairage are present in 42/230 slaughterhouses; in 9 cases in presence of workers.

### 3.2 Results from Outbreaks Questionnaires

Within the 2021-2022 time-frame, on voluntary basis we received 24 outbreak validated reports from the officers of the Preventive Departments of Local Health Units. In **Tables 5 and 6** are reported the descriptors of the outbreaks and the corrective measures taken, respectively.

**Table 5.** Descriptors of the COVID-19 Outbreaks recorded in Italian meat plants, on voluntary basis.

| Province | Species | Workers | S_C | T_W | Outbreak |  |  | Workers | Positives | % | Companies | Report |
| --- | --- | --- | --- | --- | --- | --- | --- | --- | --- | --- | --- | --- |
|  |  | (N) | (N) | (N) | dd/mm/yy |  |  | tested |  |  |  | dd/mm/yy |
|  |  |  |  |  | Start | End | Days | (N) | (N) |  | (N) |  |
| TRENTO | S (S) | 210 | 2 | 40 | 09/09/20 | 02/10/20 | 22 | 169 | 33 | 20 | 3 | 04/12/20 |
| NAPOLI | S (P) | 195 | 2 | 0 | 28/08/20 | 10/10/20 | 41 | 195 | 87 | 45 | 2 | 15/12/20 |
| TRENTO | S (S) | 206 | 3 | 71 | 08/09/20 | 23/09/20 | 15 | 140 | 33 | 24 | 4 | 05/01/21 |
| TRENTO | CP (S) | 119 | 4 | 71 | 01/09/20 | 23/09/20 | 22 | 122 | 81 | 66 | 5 | 05/01/21 |
| TRENTO | S (S) | 36 | 2 | 42 | 16/09/20 | 05/10/20 | 19 | 36 | 25 | 69 | 3 | 05/01/21 |
| TRENTO | CP (na) | 35 | 5 | 71 | 04/09/20 | 23/09/20 | 19 | 35 | 13 | 37 | 3 | 05/01/21 |
| BARI | S (C,S,SR,H) | 13 | 0 | 0 | 03/11/20 | 30/11/20 | 27 | 15 | 4 | 27 | 1 | 14/01/21 |
| BARI | S (C,S,SR,H) | 50 | 0 | 0 | 13/11/20 | 20/12/20 | 37 | 50 | 6 | 12 | 1 | 03/02/21 |
| TREVISO | S (C,H) | 19 | 5 | 26 | 20/11/20 | 27/11/20 | 17 | 18 | 0 | 0 | 1 | 12/02/21 |
| BARI | S (C,S,SR) | 438 | 7 | 71 | 23/04/20 | 19/05/20 | 26 | 487 | 112 | 23 | 7 | 26/03/21 |
| TREVISO | S (C) | 180 | 5 | 50 | 17/11/20 | 10/12/20 | 23 | 43 | 20 | 47 | 1 | 27/04/21 |
| VENEZIA | S (P) | 122 | 2 | 6 | 28/10/20 | 10/01/21 | 12 | 180 | 49 | 27 | 2 | 19/07/21 |
| RAGUSA | S (P) | 205 | 2 | na | 05/11/20 | 02/12/20 | 27 | 205 | 12 | 6 | 1 | 15/09/21 |
| RAGUSA | S (P) | 250 | 2 | na | 30/03/21 | 30/06/21 | 90 | 57 | 8 | 14 | 1 | 16/09/21 |
| MANTOVA | S (S) | 350 | 4 | 65 | 29/06/20 | 15/07/20 | 16 | 350 | 50 | 14 | 5 | 04/10/20 |
| MANTOVA | S (S) | 184 | 5 | 56 | 10/07/21 | 30/07/21 | 20 | 184 | 10 | 5 | 3 | 19/01/21 |
| MANTOVA | S, CP (S) | 350 | 2 | 60 | 10/06/21 | 25/06/21 | 15 | 123 | 38 | 31 | 2 | 30/09/21 |
| MANTOVA | CP (na) | 15 | 1 | 17 | 30/06/20 | 13/08/20 | 44 | 15 | 7 | 47 | 2 | 04/11/21 |
| MANTOVA | S, CP (C,S) | 150 | 1 | 6 | 22/02/21 | 05/04/21 | 44 | 152 | 5 | 3 | 1 | 29/09/21 |
| MANTOVA | S (S) | 270 | 3 | 40 | 16/08/21 | 24/08/21 | 8 | 8 | 1 | 13 | 3 | 20/10/21 |
| MANTOVA | CP (na) | 400 | 2 | 5 | 26/07/21 | 16/08/21 | 20 | 30 | 6 | 20 | 1 | 27/09/21 |
| MANTOVA | S (C) | 270 | 6 | 70 | 04/04/20 | 29/10/21 | 25 | 267 | 47 | 18 | 4 | 05/11/21 |
| PADOVA | S (P) | 45 | 2 | 20 | 22/10/20 | 09/11/20 | 18 | 27 | 9 | 33 | 1 | 10/08/21 |
| REGGIO C. | S (C, S, SR, H) | 7 | 0 | 0 | 01/01/21 | 30/06/21 | 180 | 7 | 7 | 100 | 1 | 26/09/22 |

2 S\_C = Subcontractors; T\_W = temporary workers; na = not available. S = Slaughterhouse; CP = processing and cutting plant; C= cattle; 3 S= Swine; P = Poultry; SR = Small Ruminants, H = horses

**Table 6.** Descriptors of the corrective measures taken in the recorded 24 outbreak management

| Measure | Plants (N) | % |
| --- | --- | --- |
| Personal Protective Equipments | 8 | 33 |
| Intensified checks about application of preventive measures | 10 | 42 |
| Cleaning and disinfection (sanitization) | 16 | 67 |
| Ventilation | 6 | 25 |
| Change in the plant structure and layout revision | 1 | 4 |
| Change in the entrance/exit procedures for external personnel | 10 | 42 |
| Change of the entrance | 10 | 42 |
| Personal Hygiene | 9 | 38 |
| Workers information | 10 | 42 |
| Increase of the working place distances | 4 | 17 |
| Physical barriers among workers and among internal vs external personnel | 4 | 17 |
| Homogeneous composition of workshift force | 1 | 4 |

## 4. Discussion

The main feature of Italian slaughterhouses and of processing/cutting plants is the rather huge number (N = 4,675 officially registered. Among them, around 20 plants have a working force above 500 up to 2,000 units: In the poultry sector, 6 industrial plants cover the 90% of the national production. Within this frame, the reduced working force in most of the plants, along to activities carried out not all the week long (**Table 2 a,b**) represent mitigation factors with respect to COVID-19 spread and related outbreaks reported in large industrial plants also as matter of higher workers density coupled also to a longer stay in working place via the use of overtime as already noted by Dyal et al., and Waltenburg et al., [3, 4].On the other side, such a diffuse presence of plants on the Italian territory, hampers a capillary monitoring activity of the preventive COVID-19 procedures in place. It’s worth noting during the pandemics (**Figure 1**) the professional resources of the Prevention Departments of the Local Health Unit in charge of the health prevention in the working places have been absorbed by other health priorities, such as contact tracing, testing, vaccination. Owing to the above, an advocacy activity based on the proposition of a Target Plan of Prevention seemed more appropriate and practicable.

The results we present in this paper do not cover all the initiatives taken to prevent and manage COVID-19 outbreaks in meat plants on the national territory. For instance, Emilia Romagna Region independently from this initiative, activated cultural mediators to inform and to form non Italian workers about procedures to be adopted to lower the risk (such as quarantine period back from holidays in the Country of origin, priority in the diagnostic tests, and then, in a second stage, support to vaccination) as reported in a Canadian – Ontario case [8]. The shared knowledge of such in a webinar organised on Sept 2020 by Istituto Superiore di Sanità with the participation of the representatives of meat plan associations and health stakeholders, contributed to the questionnaires set up.

The collected answers (N = 333) to the advocay questionnaire represent the 7% of the registered plants in Italy, and are mostly located in those districts whose Regional/Province Health Authorities agreed to activate the Target Prevention (Lombardy, N =140; Veneto, N=95), according to the flow diagram reported in **Figure 2**. The support at local level has been also determined by the high added social and economic added given to processed meat products (such as ham and salami) from such geographical areas. The reduced dimension of the meat plants in Italy may have contributed to a reduced perception of the relevance of the iniatiative, thus determining a limited participation to the survey on voluntary basis. In the United States, a survey on COVID-19 at slaughterhouses accounted for the active participation of 28 out of 50 States (56%), accounting for an average number of 3,500 meat plants, with an overall workforce of 525,000 workers [4].

Despite the 7% of adhesion to the Italian initiative, the answers recorded from the advocay questionnaire reflect the presence of those structural, environmental, and management risk factors, already reported in the literature [15, 16].

The working force management, with non permanent staff (cooperatives, third parties, autonomous workers) present in the 42% of the companies (N = 139) indicates the risk of formation of non homogeneous teams as a factor that could facilitate the contagiousness within the same workshift. This critical factor finds a feed-back in the 24 reported outbreaks, where, on average, 3 cooperatives (min/max = 0 - 6) were present (**Table 5**). In the personnel management, a combined risk indicator for the entrance and spread of COVID-19 among workers, includes the number of animals slaughtered/processed per day, the number of timeshifts per day, and the data about external workforce. Within this frame, the answers about the instrumental check of the body Temperature at the entrance recorded in the 27% of the plants only, represent another weak point of the preventive action, especially in presence of a pandemic R_t_ well >1 in the general population. The mitigation of the risk in the working place relies also on appropriate information and formation of the workers, along to the verification if the preventive measures are correctly put in place, and last but not least the activation of COVID-19 illness social valves (in our survey present in the 26% of plants, only). Again, the relevance of such factors has been highlighted by the evidences from the outbreak managements (**Table 6**), where the plant responsibles had to re-inforce the checks, re-draft the entrance and exit paths of workers and external personnel, improve information and formation (42% of the reported outbreaks), and provide devices for a better personal hygiene (in the 38% of the cases).

The overall need of a advocacy activity towards the real adoption of COVID-19 preventive measures and the check of their correct application by meat plant management is highlighted by the recorded delay in the set up of the meat plant committee formed by occupational safety responsibles and trade union representatives in charge of this task (51% of positive answers), despite the COVID-19 update of the mandatory document on the risk assessment recorded in the 83% of answers (**Figure 2**). To this respect, the recorded generation of vapour and aerosols in some working areas according to each plant structure seem a factor to be not overlooked in the update of the plant document on risk assessment. This, because it has been demonstrated Sars-CoV-2 could reach distances well over those minimum prescribed between workers (1-2 m) in presence of vapour and aerosols, as reported in the in industrial slaughterhouses, in Germany [17]. This represent a risk factor especially if vapour/areosol exposed workers do not wear adequate PPE (**Table 4**).

The ventilation, with an adequate natural air exchange represents another preventive measure stressed in the advocacy questionnaire, as far as it has been demonstrate the relative percentage of CO_2_ could be assumed as a proxy of insufficient indoor air exchange [15, 18]. When air is recycled, it is worthy to consider that a recycling percentage above 30% may be inadequate for the prevention, especially when adequate filtering systems are not in place. High percentage of recycled air may acknowledge seasonal trends, as matter of energy savings policies to keep room temperature adequate for the specific activities of the working area (chilling or air warming during summer or winter, for instance). To this purpose, as feedback, structural modifications of the ventilation have been considered in 6 out of 24 cases in the management of outbreaks (**Table 6**). Such measure has been associated to an increase of the inter-worker distance in the 17% of cases. Last but not least, in presence of a HVAC system, the air fluxes in the working area should be addressed correctly.

Considering the reported outbreaks dates, it is worthy to fix the date of July 20, as discriminant for the COVID-19 vaccination campaign extended to the Italian general population (meat plant workers included). Before such date, vaccines were administered to fragile persons and to most exposed workers categories, such those working in the healthcare settings, schools, and police forces. With respect to such date, we registered 14 outbreaks before, and 10 outbreaks later. This means vaccination could not represent alone an effective prevention in such setting, but it should accompany all the supporting preventive measures, especially when the workforce is in large part not autochtonous (Fabreau et al., 2022).

The outbreaks lasted on average 26 days (one outlier data of 180 gg excluded) (**Table 5**), with an overall 26% positivity rate among workers, in line with the prevalence reported in other papers: 3 - 24% from Waltenbur et al., [4], 18% from Di Leone et al., [13], 26% from Steinberg et al., [19], 30-40% from Vanderwaal et al., [20], 12-16% from Pokora et al., [9] 33% from Walshe et al., [15], 36% from Finci et al., [16]. Of interest, the 5 outbreaks reported from the Trentino-Alto Adige Province, where almost the same cooperatives were turned-over among the plants involved (Dario Huber, personal communication).

Among the reported outbreaks, the description of the widest Italian outbreak occurred in a poultry slaughterhouse and cutting plant (August – September 20) has been missed. From public available information as those reported on the website of the Veterinary Trade Union of the Veneto Region, positivities incolved 200/675 workers belonging to 12 different nationalities, and the massive and intensified screeneng with rapid tests allowed the 50% reduction of the activities, instead of their full withdraw (https://www.sivempveneto.it/covid-19-focolai-in-impianti-di-macellazione-il-sivemp-veneto-massima-attenzione-alla-sicurezza-dei-veterinari-ufficiali-protezioni-e-screening-costanti/).

## 5. Conclusions

To conclude, the proposition to the territory of a Target Prevention Plan on COVID-19 in meat plants, supported by guidelines and advocacy questionnaires represented a valuable tool to harmonise and risk-orient the activities of the Health Preventive Departments on the territory. The cross-checks between the results from the advocacy and from the outbreak questionnaires will help to assess and illustrate to the stakeholders the critical points to be implemented in term of preparedness, within an after-action review for such food-chain related essential work settings not linked to healthcare services. Within this frame, because COVID-19 has been acknowledged as an “emerging infectious disease of probable animal origin”, meat plants and related settings represent health, cultural, social, economic, and food safety/security environments where “One Health” preventive cost-effective approaches could be practiced.

## Data Availability

All data produced in the present study are available upon reasonable request to the authors

https://www.iss.it/documenti-in-rilievo/-/asset_publisher/btw1J82wtYzH/content/rapporto-iss-covid-19-n.-8-2021-english-version-set-up-of-a-risk-oriented-plan-for-the-control-and-management-of-covid-19-outbreaks-in-meat-plants-ad-interim-methodological-approach.-version-of-april-8-2021

## Acknowledgements

The Authors wish to thanks all the active participants to the questionnaires initiative and the followind colleagues who institutionally supported the initiative: Adelina Brusco, Silvia D’Amario, Fabio Boccuni, Bruna Maria Rondinone, Paola Tomao, Nicoletta Vonesch, and Sergio Iavicoli from INAIL; Nicoletta Cornaggia, DG Welfare Regione Lombardia; Francesca Russo, Direzione Prevenzione, Sicurezza Alimentare, Veterinaria Regione del Veneto; Mara Bernardini and Anna Padovani, Regione Emilia-Romagna; Anna Marinella Firmi and Alberto Righi, ATS Valpadana; Dario Uber, APSS Provincia di Trento.

DOI: Authors declare no competing interest

Credits

Giorgio Di Leone, Simona Savi, Gianfranco Brambilla: conceptualization and relationship with stakeholders Gianfranco Brambilla: manuscript writing and references

Flavio Napolano, Gaetano Settimo, Valerio Manno and Simona Savi: Questionnaires formulation and analysis of the answers

Domenico Lagravinese and Luigi Bertinato: manuscript supervisors

---

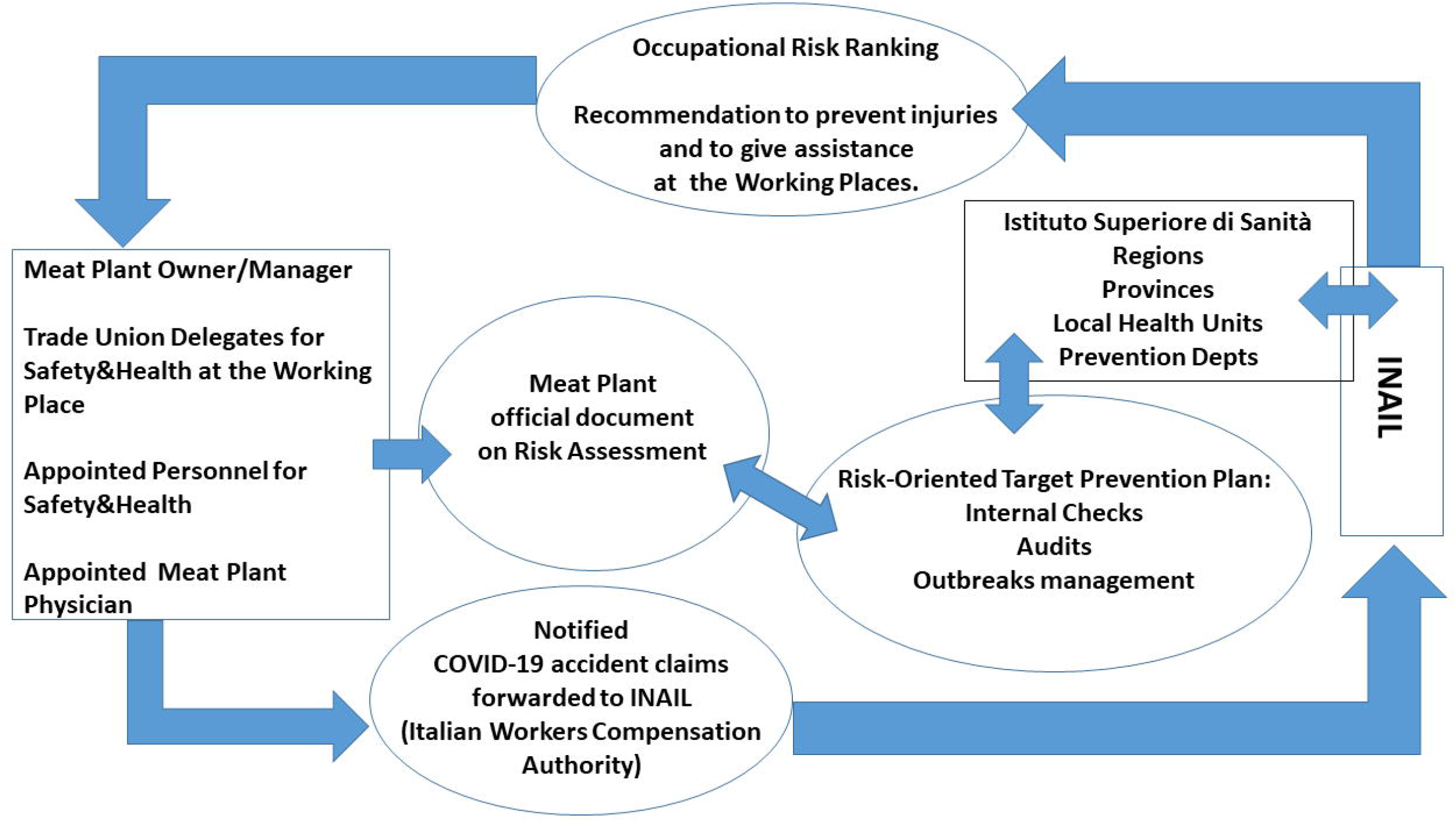

## Notes

### Competing Interest Statement

The authors have declared no competing interest.

### Funding Statement

This study did not receive any funding

